# Gut-related Immune Activation in Parkinson’s Disease with Asian *LRRK2* Risk Variants: Associations with Systemic Inflammation and Clinical Severity

**DOI:** 10.64898/2026.07.10.26357757

**Authors:** Tzi Shin Toh, Hans Xing Ding, Anis Nadhirah Khairul Anuar, Nurul Syuhada Zulhaimi, Jia Wei Hor, Yong Chiang Pang, Ing Xian Kong, Jannah Zulkefli, Yi Wen Tay, Lei Cheng Lit, Shen-Yang Lim, Ai Huey Tan

## Abstract

*LRRK2* is implicated in Parkinson’s disease (PD) microbiome–gut–brain axis. We compared plasma lipopolysaccharide-binding protein (LBP) and soluble CD14 (sCD14), markers of gut permeability and endotoxin exposure, in PD patients with/without *LRRK2* p.G2385R and/or p.R1628P, and controls, and examined their associations with systemic inflammation and clinical severity. Neither marker differed between groups. Across PD patients, LBP correlated with higher IL-6, TNF-α and worse motor function, while sCD14 correlated with higher IL-6, CCL5 and worse constipation. These findings highlight the clinical relevance of endotoxin-related immune signaling in PD, without *LRRK2* risk variant-specific associations and identify LBP as an emerging marker of inflammatory burden.

## INTRODUCTION

Intestinal barrier dysfunction and gut inflammation are key components of the microbiome– gut–brain axis in Parkinson’s disease (PD).^1-3^ Gut microbiome alterations in PD have been widely reported,^4-6^ with consistent alterations in several Gram-negative taxa (e.g., *Akkermansia, Megasphaera, Desulfovibrio, Escherichia*), raising the possibility of altered lipopolysaccharide (LPS) exposure, a potent trigger of local and systemic inflammation.^1,7^

Increased intestinal permeability may promote LPS translocation from the gut lumen into the systemic circulation, where LPS binds to LPS-binding protein (LBP), facilitating its transfer to cluster of differentiation 14 (CD14) and the Toll-like receptor 4 (TLR4)/myeloid differentiation protein 2 (MD2) complex, thereby initiating innate immune signaling.^7,8^ LPS engagement of membrane-bound CD14 also induces the release of soluble CD14 (sCD14). Accordingly, LBP is widely used as a surrogate marker of gut permeability and endotoxin exposure, whereas sCD14 reflects host immune activation following endotoxin exposure.^9,10^ Previous studies have assessed intestinal barrier dysfunction and gut inflammation using calprotectin,^11-18^ zonulin,^11,12,14-19^ and LBP.^7,15,20-26^ Meanwhile, evidence for sCD14 in PD remains limited to two small studies (n=19–40), both of which measured sCD14 alongside LBP but reported conflicting results.^21,25^ Importantly, whether these gut-related immune markers are associated with systemic inflammation and clinical severity in PD remains largely unexplored.

Host genetic factors are increasingly recognized as important modulators of the gut microbiome and immune responses. Among these, *LRRK2* has emerged as a compelling candidate linking genetic susceptibility, intestinal inflammation, and PD.^27,28^ Besides being a common genetic contributors to PD, *LRRK2* is a susceptibility gene for Crohn’s disease, and epidemiological studies have reported increased PD risk among individuals with inflammatory bowel disease.^27,28^ Experimental studies have demonstrated that LRRK2-mediated immune signaling promotes intestinal inflammation and neuroinflammation.^29-31^ Despite these experimental observation, it remains unclear whether *LRRK2* genotype influences gut permeability and endotoxin translocation-related immune activation in PD patients. This is particularly relevant in Asian populations, where the common p.G2385R and p.R1628P risk variants each account for ∼5–10% of PD cases.^32^

Therefore, we compared circulating LBP and sCD14 concentrations in PD patients with and without *LRRK2* p.G2385R and/or p.R1628P variants and healthy controls (HC), and examined their associations with systemic inflammation and clinical characteristics in PD.

## METHODS

### Participant Recruitment and Clinical Evaluation

Manifesting carriers of the *LRRK2* p.G2385R variant (PD-G2385R; n=56), p.R1628P variant (PD-R1628P; n=60), both variants (PD-G2385R+R1628P; n=5), patients with idiopathic PD (iPD; n=61), and HC (n=58) from a previous study were included.^33^ Patients were diagnosed by movement disorder specialists according to standard clinical diagnostic criteria as previously described.^33,34^ Clinical assessments included International Parkinson and Movement Disorder Society-Unified Parkinson’s Disease Rating Scale (MDS-UPDRS), Clinical Impression of Severity Index for Parkinson’s Disease (CISI-PD), Montreal Cognitive Assessment (MoCA), and constipation measures (Bristol Stool Form Scale, ROME IV criteria, Patient Assessment of Constipation-Symptoms [PAC-SYM]).

### Measurements of Plasma LBP, sCD14, Chemokines, and Cytokines

Ten milliliters of peripheral blood were collected into EDTA tubes and centrifuged within 30 minutes of collection. Plasma was aliquoted and stored at –80°C until analysis. Plasma LBP, sCD14, chemokines (CCL2, CCL5, CX3CL1, VCAM1), and cytokines (IL-6, TNF-α) were measured using the ELLA™ multiplex immunoassay system (ProteinSimple, California, USA) according to the manufacturer’s instructions.

### Statistical Analysis

Statistical analyses were performed using R (v4.6.0). Data normality was assessed using the Shapiro-Wilk test. Between-group differences were evaluated using analysis of variance, Kruskal-Wallis, or chi-square tests as appropriate. Spearman’s correlation analysis was used to assess associations of LBP and sCD14 with systemic inflammatory markers and clinical variables. Generalized linear models were used to examine these associations while adjusting for potential covariates.

## RESULTS

No significant between-group differences were observed in age, sex, body mass index, and comorbidities (**Table 1**). Plasma LBP and sCD14 levels did not differ significantly among HC, iPD, PD-G2385R, PD-R1628P, and double-variant carriers (p=0.172 and 0.326, respectively; **Figure 1A**). However, LBP and sCD14 were positively correlated in the overall cohort (r_s_=0.418, p<0.001), overall PD group (r_s_=0.420, p<0.001), and within iPD (r_s_=0.476, p<0.001), PD-G2385R (r_s_=0.352, p=0.008), and PD-R1628P (r_s_=0.383, p=0.003). These associations remained significant after adjustment for age (all p<0.001).

**Table 1:**
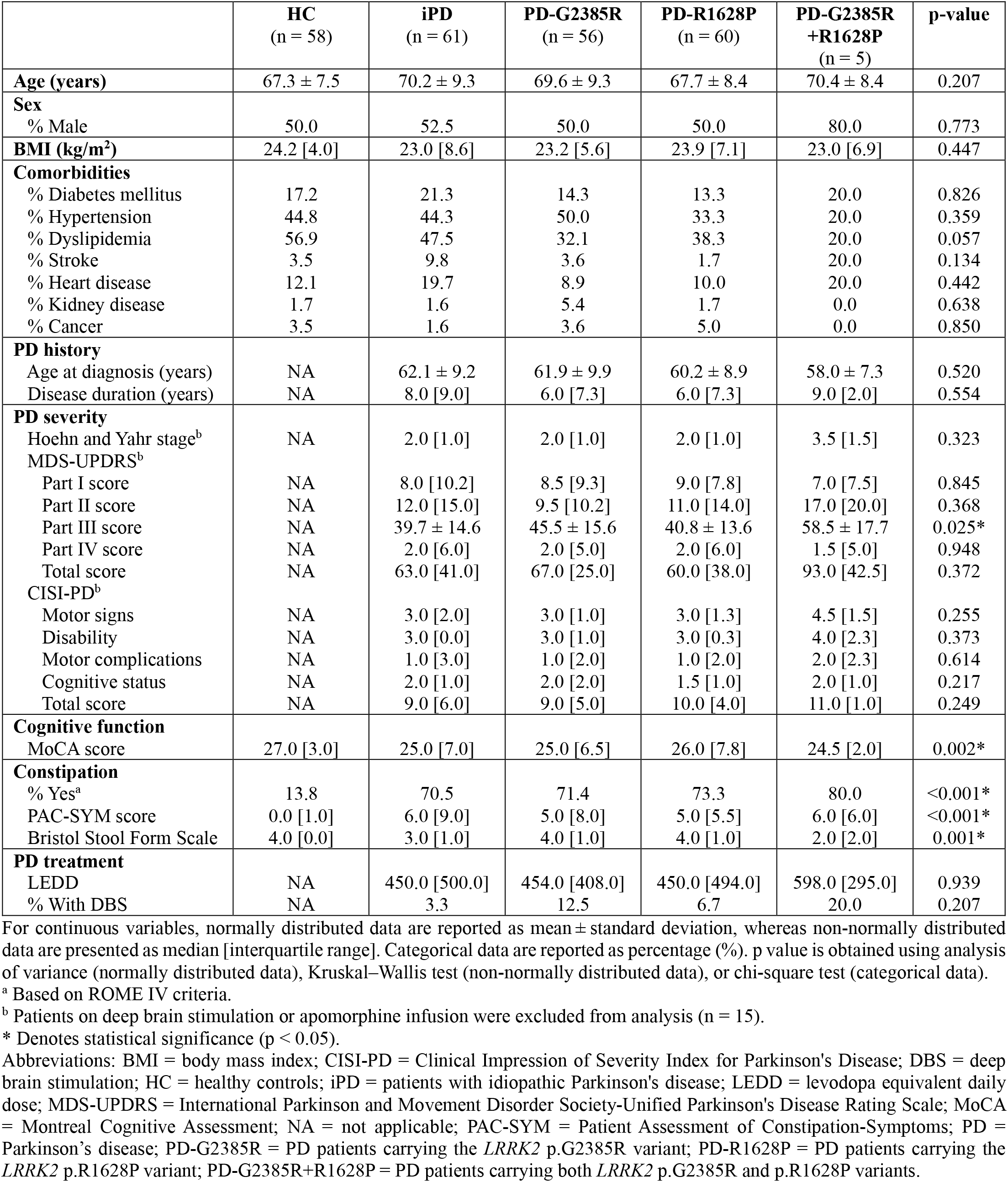
Clinico-demographic characteristics of all participants in the study.

**Figure 1:**
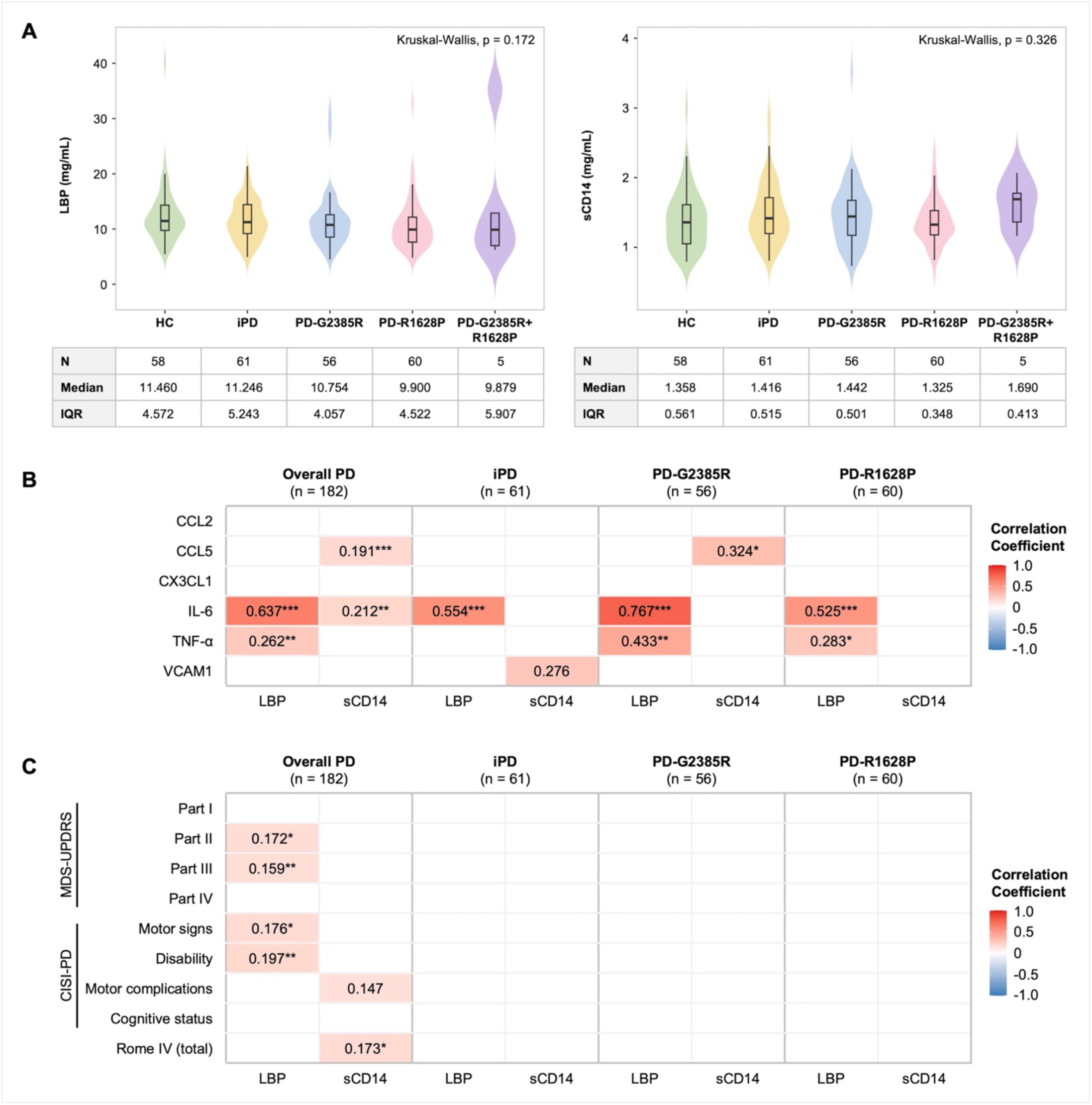
Plasma levels of LBP and sCD14 and their correlations with systemic inflammatory markers and clinical variables. (**A**) Comparison of plasma LBP and sCD14 levels among HC, iPD, PD-G2385R, PD-R1628P, and double-variant carriers. (**B**) Correlations of plasma LBP and sCD14 levels with systemic inflammatory markers in the overall PD cohort and in each PD subgroup. Colored cells indicate significant correlations (Spearman’s rank correlation, p < 0.05). Asterisks denote correlations that remained significant after adjustment for age using GLM (*** p < 0.001, ** p < 0.01, * p < 0.05). (**C**) Correlations of plasma LBP and sCD14 with clinical variables in the overall PD cohort and in each PD subgroup. Colored cells indicate significant correlations (Spearman’s rank correlation, p < 0.05). Asterisks denote correlations that remained significant after covariate adjustment using GLM (*** p < 0.001, ** p < 0.01, * p < 0.05). Analyses of MDS-UPDRS Parts I-III, CISI-PD motor signs, and CISI-PD disability were adjusted for age and disease duration. Analyses of MDS-UPDRS Part IV and CISI-PD motor complications were adjusted for age at diagnosis and disease duration. Analyses of MoCA and CISI-PD cognitive status were adjusted for age.

Plasma LBP levels were positively correlated with IL-6 in the overall PD cohort (r_s_=0.637), and within iPD (r_s_=0.554), PD-G2385R (r_s_=0.767), and PD-R1628P (r_s_=0.525) (all p<0.001; **Figure 1B**). Weaker but significant correlations between LBP and TNF-α were observed in the overall PD cohort (r_s_=0.262, p=0.001), PD-G2385R (r_s_=0.433, p=0.001) and PD-R1628P (r_s_=0.283, p=0.031). In contrast, sCD14 showed less consistent associations. Significant correlations were observed with CCL5 in the overall PD cohort (r_s_=0.191, p=0.012) and PD-G2385R (r_s_=0.324, p=0.019), with IL-6 in the overall PD cohort (r_s_=0.212, p=0.005), and with VCAM1 in iPD (r_s_=0.276, p=0.040). All associations remained significant after adjustment for age (p<0.05), except for sCD14 and VCAM1 in iPD.

In the overall PD cohort, higher LBP levels were correlated with worse motor function (higher MDS-UPDRS Part III [r_s_=0.159, p=0.033] and CISI-PD motor signs scores [r_s_=0.176, p=0.018]) and disability (higher MDS-UPDRS Part II [r_s_=0.172, p=0.022] and CISI-PD disability scores [r_s_=0.197, p=0.008]) (**Figure 1C**). Higher sCD14 levels were associated with worse motor complications (higher CISI-PD motor complications scores [r_s_=0.147, p=0.048]) and constipation severity (higher ROME IV scores [r_s_=0.173, p=0.020]). After covariate adjustment, all associations remained significant, except for the association between sCD14 and CISI-PD motor complications scores. No associations were observed with MoCA, PAC-SYM or Bristol Stool Form Scale.

## DISCUSSION

This study provides the first evaluation of circulating LBP and sCD14 in PD stratified by the Asian-prevalent *LRRK2* p.G2385R and p.R1628P risk variants. Although neither marker differed between groups, higher LBP levels were correlated with a more proinflammatory profile (higher IL-6 and TNF-α levels), and worse motor function and disability in the overall PD cohort. In contrast, sCD14 showed modest and inconsistent associations, with higher levels associated with higher plasma IL-6 and CCL5 and worse constipation in the overall PD cohort. These findings suggest that gut permeability and endotoxin-related immune activity may be linked to systemic inflammation with clinical consequences in PD, irrespective of *LRRK2* risk variant status, with plasma LBP emerging as an informative marker of gut permeability and endotoxin exposure.

LBP showed robust correlations with systemic inflammatory markers and clinical severity in PD. Previous studies (n=10–397) have reported inconsistent plasma LBP findings in PD compared to HC, with most reporting lower levels in PD,^7,20,21,24-26^ while others observed higher or comparable levels.^15,22^ Mechanistically, LBP facilitates LPS transfer to the CD14/TLR4 receptor complex, promoting innate immune signaling and downstream release of proinflammatory cytokines, including IL-6 and TNF-α.^7^ Elevated IL-6 has been linked to faster clinical progression,^35,36^ while TNF-α has been implicated in intestinal inflammation, oxidative stress, and dopaminergic neurodegeneration in PD.^37,38^ Besides reports of higher LBP in advanced versus early PD,^23^ several studies have demonstrated positive associations between plasma LBP levels with worse motor impairment and constipation in PD.^20,24^ Consistent with these findings, higher LBP levels in our cohort correlated with increased IL-6 and TNF-α concentrations, as well as greater motor impairment and disability. Collectively, these findings support gut-related endotoxin exposure as a potential contributor to systemic inflammation and PD severity. Despite the role of LRRK2 in immune regulation, p.G2385R and p.R1628P variants did not substantially modify these associations, suggesting that other genetic or environmental factors may shape gut–immune interactions in PD. The consistent associations between LBP, systemic inflammation, and clinical severity highlight its potential utility as a marker for patient stratification and disease prognosis, particularly in the context of future gut-targeted therapeutic trials, warranting validation in prospective longitudinal studies.

In contrast, sCD14 showed weaker associations with systemic inflammatory markers and clinical features, with significant correlations observed only in the overall PD cohort. Its modest correlations with IL-6 and CCL5 suggest that circulating sCD14 may reflect only a limited component of endotoxin-induced immune activation, possibly related to monocyte activation. Although not observed in our cohort, a previous investigation reported higher sCD14 levels in PD despite similar LPS concentrations between PD and controls.^25^ These findings suggest that sCD14 may represent a marker of host immune activation to endotoxin exposure rather than a direct reflection of circulating LPS burden.^25^ Further studies are warranted to clarify its role in gut-related immune dysregulation in PD.

This study has several limitations. The cross-sectional design precludes causal inference. Circulating LBP and sCD14 are indirect markers that do not directly measure intestinal permeability, microbial translocation, or endotoxin exposure. The systemic inflammatory panel was limited to selected cytokines and chemokines and may not capture the full systemic inflammatory spectrum. Gut microbiome profiling was not included, limiting assessment of microbial contributors. Future longitudinal and in vivo studies incorporating direct endotoxin measurements, microbiome profiling, and genetic characterization are needed to clarify gut-related immune mechanisms in PD.

## CONCLUSION

Plasma LBP and sCD14 levels did not differ between HC, iPD, and PD patients carrying Asian-prevalent *LRRK2* p.G2385R and p.R1628P risk variants. Nevertheless, LBP demonstrated robust associations with systemic inflammation and motor impairment, whereas sCD14 showed more modest associations with inflammatory markers and constipation severity. These findings support a role for gut-related endotoxin-induced immune activation in PD that is not substantially modified by these *LRRK2* risk variants. Plasma LBP may represent a promising marker of inflammatory burden for patient stratification and disease prognosis, warranting validation in prospective longitudinal studies.

## ACKNOWLEDGEMENTS

We thank Samantha Hutten and Shalini Padmanabhan from the Michael J. Fox Foundation for Parkinson’s Research for their kind assistance with project and grant management. We also thank Associate Professor Dr Reena Rajasuriar and the Immunotherapeutics Laboratory at the Faculty of Medicine, University of Malaya, for the laboratory assistance and support in this work. We are sincerely grateful to all research participants and their families and caregivers for their invaluable contributions to this study, without which this work would not have been possible.

## AUTHOR CONTRIBUTIONS

AHT conceptualized and designed the study. TST, HXD, ANKA, NSZ, JWH, YCP, IXK, JZ, YWT, LCL, SYL, and AHT contributed to data acquisition. TST and AHT analyzed and interpreted the data. TST wrote the first draft of the manuscript, and all authors contributed to its revision.

## DATA AVAILABILITY

The data supporting the findings of this study are available from the corresponding author on reasonable request but are not publicly available due to privacy and ethical restrictions.

